# Sampling and host-strain interactions shape detection of *Staphylococcus aureus* transmission in hospitals

**DOI:** 10.64898/2026.04.28.26351952

**Authors:** Kristine B. Rabii, Courtney Takats, Gregory Putzel, Alice Tillman, Magdalena Podkowik, Julian McWilliams, Nora Samhadaneh, Julia Shenderovich, Natalia Arguelles, Anusha Srivastava, Karl Drlica, Victor J. Torres, Alejandro Pironti, Sarah E. Hochman, Audrey Renson, Bo Shopsin

## Abstract

In principle, whole-genome sequencing of *Staphylococcus aureus* in hospitals is most effective when used prospectively, but its practical limits are unclear. We performed large-scale genomic surveillance across two interconnected hospitals, sequencing 4,779 *S. aureus* isolates from admission screening and clinical cultures. Integration of genomic and epidemiologic data identified 361 transmission events undetected by standard surveillance. Despite dense sampling, most events (90%) were detected only at readmission, indicating that transmission escapes recognition during the index hospitalization. Serial or discharge screening is likely required to capture transmission. Detection depended on sampling: clinical isolates alone were insufficient, and nearly all events required screening and repeated sampling. Because such approaches are unlikely to scale if applied universally, surveillance must be targeted. Transmission concentrated in methicillin-resistant strains, and it increased when healthcare exposure aligned with hospital-associated strain lineage. These findings help define the limits of prospective genomic surveillance and provide a framework for targeted detection.

**Main point:** This work indicates that most MRSA transmission arises from asymptomatic carriers and is detected only after readmission, highlighting the need for targeted serial or discharge screening of high-risk patients to identify and interrupt hospital transmission.

## INTRODUCTION

*Staphylococcus aureus*, especially methicillin-resistant strains (MRSA), is a leading cause of morbidity and mortality among hospitalized patients worldwide [1–5]. MRSA is notorious for causing outbreaks in healthcare settings [6, 7], and hospital transmission is an upstream driver of infection risk [8]. Accordingly, real-time identification of transmission events has important implications for infection control, enabling timely intervention to limit spread and reduce infection burden.

Epidemiological surveillance often detects outbreaks only after they have spread to numerous patients, largely because it cannot resolve the genetic relatedness among strains. Genomic approaches address this limitation but are typically used retrospectively to investigate hospital MRSA outbreaks already identified by epidemiological methods [6, 7]. Whole-genome sequencing (WGS) is likely most effective when used prospectively as part of genomic surveillance [9–13]. Routine sequencing, rather than analysis restricted to suspected outbreaks, would enable detection of previously unnoticed transmission clusters and early intervention before outbreaks become established. This proactive approach is particularly valuable in urban health systems, where MRSA is endemic and its high prevalence limits the effectiveness of traditional epidemiologic methods for detecting outbreaks, let alone individual transmission events.

Most genomic surveillance relies on passive case-finding from infrequent infections such as bacteremia, even though the majority of transmissions likely originate from asymptomatic carriers [14, 15]. Consequently, many transmissions are detected months after they occur, often during hospital readmission [10, 15]. Denser sampling of colonizing isolates and inclusion of non-blood clinical isolates across wards, coupled with resampling at the time of subsequent admissions, may move real-time detection beyond the sparse signal provided by sequencing only bloodstream isolates.

To test this idea, we initiated a genomic surveillance program across two New York City hospitals, combining active screening for asymptomatic *S. aureus* colonization with WGS of colonizing isolates, clinical isolates from infections, and spatiotemporal analysis of related isolates. Over 14 months, we identified 361 endemic transmission events undetected by standard epidemiologic methods, which detect outbreaks but not endemic transmission; epidemiologic surveillance identified no outbreaks. However, even with dense sampling, transmission was rarely detected in real time, underscoring a fundamental limitation of surveillance. Nonetheless, transmission rate increased when healthcare exposure aligned with MRSA, particularly hospital strains, consistent with host characteristics and strain lineage jointly shaping transmission risk. This finding suggests a host-pathogen relationship that can help guide precision genomic surveillance toward patients and strains most likely to drive transmission.

## METHODS

### Study Design

We conducted a 14-month observational cohort study (October 1, 2022-December 1, 2023) in two interconnected hospitals in Brooklyn and Manhattan, New York. Adult (≥18) patients admitted to Medicine, Hematology/Oncology, Transplant (solid organ and hematopoietic stem cell) and intensive care units underwent routine nasal swab screening at admission for culture-based identification of methicillin-susceptible *S. aureus* (MSSA) and MRSA. All isolates from colonization screening were whole-genome sequenced. Among clinical isolates, all MRSA isolates (e.g., blood, sputum) were sequenced, whereas MSSA sequencing was limited to blood culture isolates due to volume and cost constraints. This approach reflects prioritization of MRSA surveillance and pilot data indicating that colonization is a major source of transmission. From a subset of patients, multiple isolates were sequenced from a subset of patients with repeat hospitalizations, surveillance cultures, or serial clinical cultures. Isolates were sub-cultured in the clinical microbiology laboratory and transferred to the research laboratory for genomic analysis. Patients with *S. aureus* colonization underwent routine decolonization per institutional protocols (Supplementary Methods). The study was approved by the NYULH Institutional Review Board (s24-01872).

### Whole-genome Sequencing and Quality Control

Genomic DNA was extracted from *S. aureus* isolates using a KingFisher Flex automated instrument (Thermo Fisher) and the MagMAX DNA Multi-Sample Ultra 2.0 kit (Applied Biosystems). De-identified isolate and metadata were imported from a central REDCap repository [16]. DNA libraries were sequenced on an Illumina NovaSeq instrument, generating paired-end 150 bp reads. Reads were filtered, trimmed, and adapter-stripped using fastp version 0.20 [17] and then assembled using Unicycler version 0.4.8 in conservative mode [18]. Cleaned reads were mapped to their assemblies using BWA 0.7.17 [19] to determine coverage; isolates with mean depth below 100 were excluded (retained isolates: ∼120–520x; N50 = 46– 847 kbp). Assemblies were taxonomically classified using GTDBTK v1.5.1 (GTDB release 202) [20], and only assemblies identified as *S. aureus* were retained. To identify mixed-strain contamination, reads were mapped to CC8 (FPR3757; NCBI ID: GCF_000013465.1) and CC5 (JH1; NCBI ID: GCA_000017125.1) reference genomes using Snippy v4.6.0 [21]. Isolates excessive heterozygosity in the alignment (>3,500 sites vs FPR3757 or >4,000 vs JH1; *n* = 258) or > 10% contamination predicted by ConFindr v0.7.4 [22] were excluded. Details of genotyping, variant calling, and phylogenetic analysis are provided in Supplementary Methods.

### Transmission Detection

Transmission of *S. aureus* was estimated using a threshold of 20 single-nucleotide variants (SNVs) between patients (Supplementary Methods). Isolates differing by ≤20 SNVs were evaluated for epidemiologic linkage using temporal and spatial data from electronic health records, including admissions, discharges, ward movements, procedures, and provider contacts. Transmission events were defined as patient pairs with isolates differing by ≤20 SNVs and epidemiologic linkage, either direct (shared unit, room, provider, or procedure with overlapping admissions) or indirect (shared ward [excluding the emergency department] within six months without overlap) [10, 23]. Events separated by >6 months were excluded, since 99% of epidemiologically linked pairs occurred within this interval. Transmission clusters were constructed from linked patient pairs (Supplementary Methods).

Transmission events were classified as real-time or delayed based on timing of isolate collection relative to hospitalization. Real-time detection was defined as genetically linked isolates collected during the same admission while both patients were hospitalized. Delayed detection was defined as genetically linked isolates identified during subsequent admissions, even when epidemiologic linkage occurred previously (Figure S1).

### Host–pathogen Interactions in Transmission: Modeling Healthcare Dependence

To assess whether transmission varied with alignment between host characteristics and strain lineage, we classified isolates as hospital-associated (CC5 MRSA) or community-associated (CC8 MRSA), with MSSA as a comparator. We derived a composite healthcare dependence (HD) score capturing healthcare exposure and comorbidity from demographic, clinical, and utilization variables, defined as the predicted probability from a logistic regression model of CC5 transmission using age, sex, Charlson score, smoking status, housing (home vs. skilled nursing facility), and cumulative healthcare utilization in the prior 6 months (hospital days, antibiotics, airway devices, central venous and peripherally inserted catheters, and drains). Age was modeled with a 3-degree-of-freedom natural cubic spline; other variables were linear. Missing smoking and housing data were multiply imputed using chained equations. The HD score showed good separation across covariates (Supplementary Table 1).

We modeled transmission probability as a function of HD and strain lineage using generalized estimating equations, specifying HD as a 3-degree-of-freedom natural cubic spline with interaction by strain type (CC5, CC8, MSSA) and accounting for clustering by patient. Results are presented as predicted probabilities with 95% confidence intervals, and differences in HD–transmission relationships across lineages were assessed using a omnibus test of spline interaction terms.

## RESULTS

### Study Participants and Isolate Characteristics

We genome-sequenced 4,779 *S. aureus* isolates from 3,550 hospitalized adults (≥18 years) admitted between October 1, 2022 and December 1, 2023. During this 14-month period, there were 98,506 adult admissions involving 305,673 ward-to-ward transfers across the two hospitals. Median patient age was 66.5 years [18-102, interquartile range (IQR) 50-77]. Among the isolates, 79% (3,777 from 3,196 patients) were obtained from colonization screening cultures, and 21% (1,002 from 534 patients) from clinical cultures. Approximately two-thirds (67%) were admitted to the Manhattan hospital and one-third to the Brooklyn hospital, representing tertiary and community-based academic hospitals, respectively. Among *S. aureus*-positive surveillance screening cultures, 28.5% (1,078/3,777) were MRSA-positive. Overall, 64% (1,078/1,677) of MRSA isolates were identified from screening and 36% (599/1,677) from clinical cultures, including 44% (262/599) from blood. Among all *S. aureus* blood cultures (*n* = 663), 40% (262/663) were MRSA.

Whole-genome sequencing identified clonal complex (CC) 8 as the predominant MRSA lineage (*n* = 891 [53%]), followed by CC5 (*n* = 544 [33%]), with the remainder comprising other clonal complexes (Figure 1A). Within CC8, strains partitioned between the USA300 (76%) and USA500 (18%) lineages, whereas CC5 was dominated by the healthcare-associated USA100 (48%) and USA800 (51%) lineages (Figure 1B). Thus, MRSA was concentrated in CC5 and CC8, the two dominant lineages in New York City and the United States [24–26].

**Figure 1.**
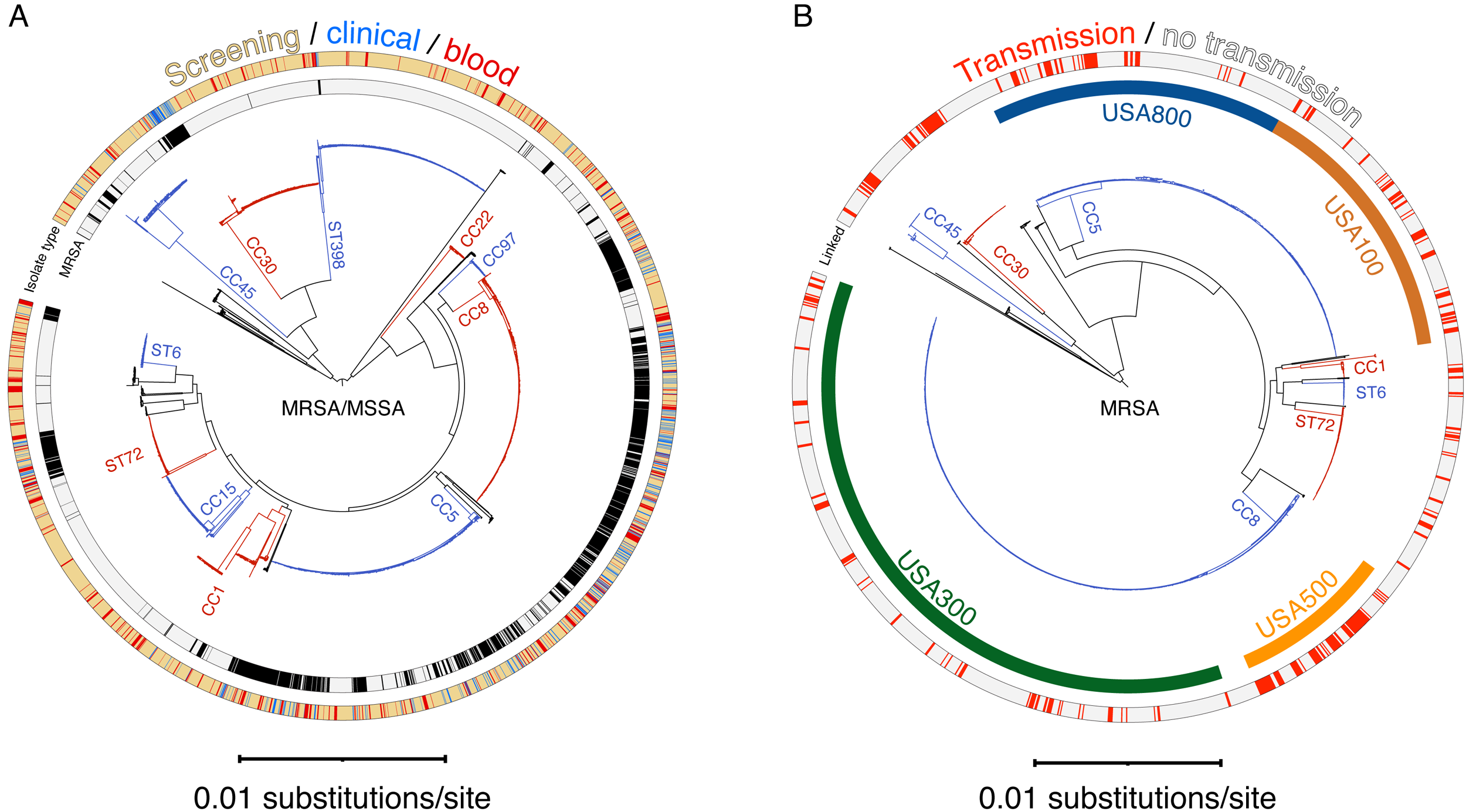
Phylogeny of *S. aureus* isolates and mapping of MRSA transmission events. **(A)** Maximum likelihood phylogeny of all isolates (*n* = 4,783 plus reference [CC8, FPR3757]). The outer ring denotes sample source (tan, screening/colonization; red, blood culture; blue, other clinical isolates). The inner ring indicates methicillin resistance (MRSA, black; MSSA, grey). MLST-defined clonal complexes (CC) are labeled directly on the phylogenetic branches. **(B)** Maximum likelihood phylogeny of MRSA isolates (*n* = 1,648 plus reference). The outer ring denotes transmission classification (red, transmission; grey, no transmission), defined by combined genomic relatedness (≤20 SNVs) and epidemiologic linkage, including both direct and indirect links (Methods). CC are labeled directly on the phylogenetic branches. The inner ring indicates lineage annotations for CC5 (USA100, USA800; defined as CC5 with SCC*mec* II or IV, respectively) and CC8 (USA300, USA500; defined as CC8 with Panton–Valentine leukocidin, arginine catabolic mobile element, and SCC*mec* IV for USA300, and SCC*mec* IV for USA500). Lineage assignments were based on the presence of defining elements in >70% of isolates within each CC, identified using in silico genomic detection, and are color-coded.

Analysis of MSSA isolates (65%; *n* = 3,102) focused on colonizing isolates obtained through surveillance to detect cryptic transmission. MSSA clinical isolate sequencing was restricted to blood isolates due to volume and cost constraints (Methods). ST398 (*n* = 586 [19%]), CC5 (*n* = 522 [17%]), and CC8 (*n* = 440 [14%]) were the most common MSSA lineages, followed by CC30, CC15, and CC1.

### Hospital Transmission of *S. aureus*

Genome sequencing identified 630 genomically linked isolate pairs involving 444 patients (12.5% of 3,550 patients; Table 1) whose genetic distances were consistent with recent transmission (≤20 single-nucleotide variants [SNVs]; Supplementary Methods). Because isolates, and thus patients, could participate in multiple pairwise links, the number of pairs exceeds the number of unique isolates. These links grouped patients into 162 genomic clusters, defined as groups of ≥2 patients connected by chains of isolates differing by ≤20 SNVs. Cluster size was typically small (mean 2.7 patients), although certain lineages (e.g., MRSA CC30) produced larger clusters (mean 11.5 patients; maximum 20). Thus, genomic links were common, but clustering was generally limited in extent.

**Table 1.**
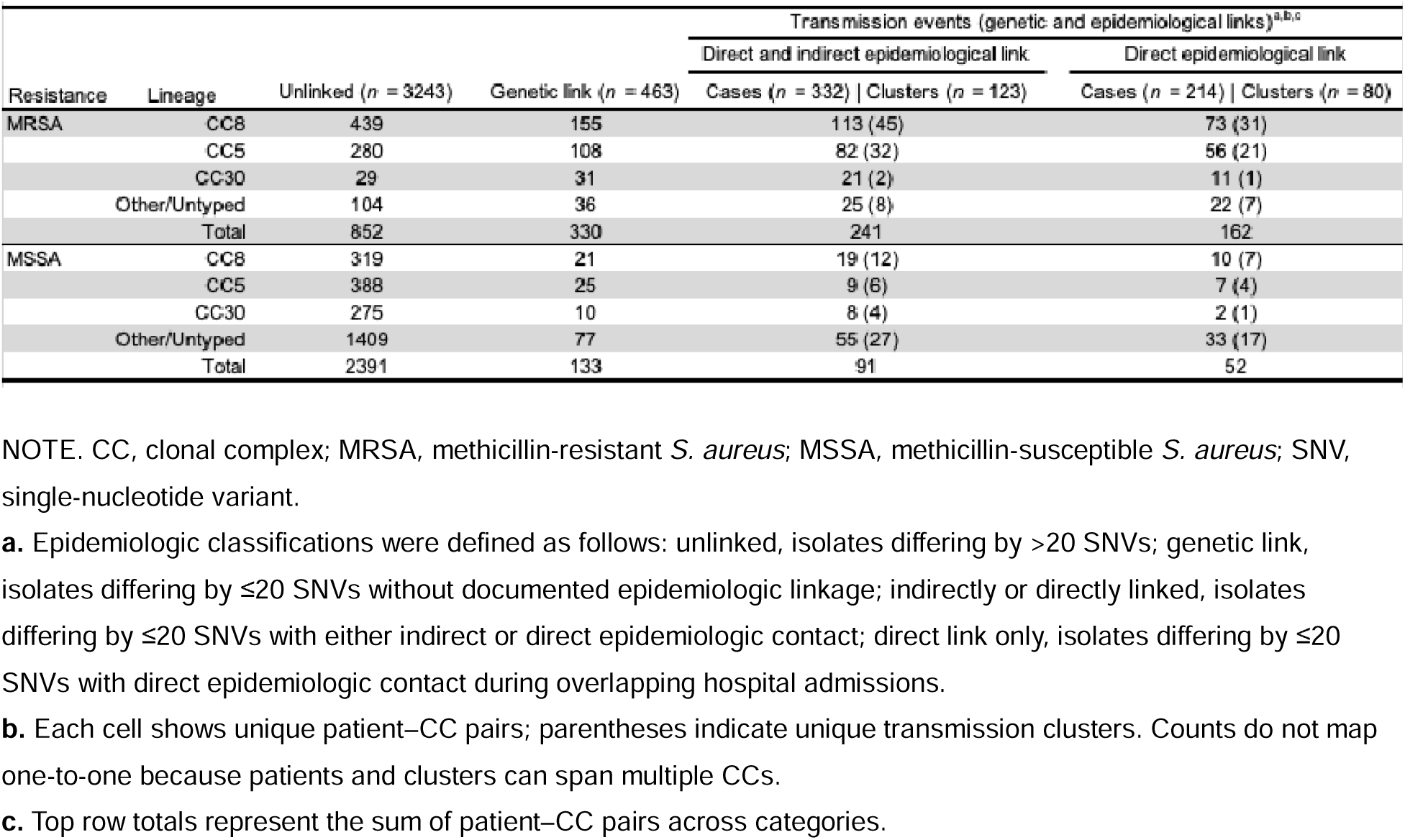
Genomic and epidemiologic classification of isolates by methicillin resistance and clonal complex. NOTE. CC, clonal complex; MRSA, methicillin-resistant *S. aureus*; MSSA, methicillin-susceptible *S. aureus*; SNV, single-nucleotide variant. **a.** Epidemiologic classifications were defined as follows: unlinked, isolates differing by >20 SNVs; genetic link, isolates differing by ≤20 SNVs without documented epidemiologic linkage; indirectly or directly linked, isolates differing by ≤20 SNVs with either indirect or direct epidemiologic contact; direct link only, isolates differing by ≤20 SNVs with direct epidemiologic contact during overlapping hospital admissions. **b.** Each cell shows unique patient–CC pairs; parentheses indicate unique transmission clusters. Counts do not map one-to-one because patients and clusters can span multiple CCs. **c.** Top row totals represent the sum of patient–CC pairs across categories.

### Genomic Transmission Thresholds

Using epidemiologic linkage based on shared ward exposure (see below), most patients with genomically linked isolates (332 of 463, 72%) had identifiable epidemiologic connections and were classified as involved in transmission. Because this definition prioritizes specificity, some genomically linked cases lacking identifiable ward contact (28%) may represent true in-hospital transmission through unobserved intermediates (false negatives), whereas others may reflect transmission outside the hospital. Consistent with this stringency, unlinked isolates (>20 SNVs) rarely shared ward contact, indicating a low false-positive rate of the epidemiologic definition (i.e., few instances in which patients without genomic evidence of transmission were nonetheless classified as epidemiologically linked). Among unrelated isolate pairs, shared ward contact occurred in <2.4% of cases.

Consistent with this, epidemiologic linkage was strongly associated with genetic relatedness. Pairing each case with its closest genetic match showed a direct relationship between genetic distance and ward contact (Figure S2), as expected [10, 23], with ∼25 SNVs capturing 95% of epidemiologically supported events. A complementary approach using within-host diversity and estimated mutation rates (Figure S3) [23] yielded a similar estimate (18.5 SNVs), based on the upper bound of within-host diversity (13.4 SNVs) plus expected mutations over six months (2 × 2.5 SNVs). Together, these findings indicate that a ≤20-SNV threshold captures most epidemiologically supported transmission events, yields few links among genetically unrelated isolates (>20 SNVs), and is consistent with within-host diversity and expected evolutionary rates ([10, 23]; Supplementary Methods).

### Integration of Genomic and Epidemiological Data

We next evaluated epidemiologic relatedness based on shared ward exposure over time (direct overlap, indirect contact, or no contact; Methods). Genomically linked pairs were classified as directly or indirectly linked, with direct links presented separately to highlight the most stringently supported transmission events (Table 1). When isolates from a patient were linked to multiple others, classification was assigned by the strongest epidemiologic connection, with direct links superseding indirect. This analysis identified 332 patients with direct or indirect epidemiologic connections, forming 123 transmission clusters; none were detected by routine epidemiologic surveillance, indicating that cryptic in-hospital transmission is common. Because genomic studies often analyze a single isolate per patient (e.g., first available [14]), we assessed the impact of restricting analysis to one isolate per patient. Restricting to the first isolate would have missed 33 of 143 direct transmission events (23%). In most cases (86%), the additional isolate belonged to a different clonal complex than the initial isolate, consistent with strain replacement or co-colonization rather than within-host diversification. Thus, repeated sampling (surveillance on subsequent admissions and/or clinical samples during hospitalization) was essential to capture transmission involving previously unsampled strains. Although clinical isolates were more likely to transmit once present (19% vs 12.6%), nearly all transmission events (∼97%) involved at least one colonizing isolate, underscoring the central role of asymptomatic carriage.

### Transmission by Clone and Lineage

We analyzed methicillin susceptibility and clonal complex distribution at the patient and cluster levels, stratified by epidemiologic classification (Figure 1B, Table 1). MSSA comprised most sequenced isolates and predominated among genetically unlinked cases, whereas MRSA accounted for nearly threefold more transmission events than MSSA (330/463, 74%), a pattern that persisted when restricted to directly linked transmissions (169/214, 76%). Additionally, CC5 and CC8 accounted for the largest share of direct and indirect links; among directly linked MRSA clusters, 45% were CC8 (73/162). Because the dataset included a broader range of clinical isolates for MRSA than MSSA, we performed a sensitivity analysis restricted to bloodstream isolates sampled in both groups. In this subset, MRSA accounted for 75% (27/36) of transmission events, with CC8 responsible for 36% (13/36). Thus, MRSA, particularly CC5 and CC8, disproportionately drives in-hospital transmission, with CC8 contributing the greater overall burden (113 vs 82 genetically linked patients; 108 vs 84 cases) while showing a similar proportional likelihood of transmission to CC5 (∼32% vs ∼24% of transmission pairs).

Transmission cluster size (integrating genomic and epidemiologic data) varied by lineage: CC5 and CC8 clusters were typically small (∼2.4 patients on average), whereas CC30 clusters, though rare, were larger (∼5.5 patients) across both MRSA and MSSA. Additionally, 46 patients (3%) carried isolates from more than one clonal complex; 37% of these were linked to a transmission cluster. Thus, clustering is shaped by both resistance and lineage, and patients within larger clusters may act as transmission hubs, sometimes acquiring multiple clones.

### Detection of Transmission Events: Real-time and Delayed

Genomic studies of bloodstream isolates often detect *S. aureus* transmission only retrospectively [15], likely because transmission originates from asymptomatic carriers long before infection. A UK study that included clinical and colonization isolates was a step forward but it did not quantify this gap [10]. We addressed this by measuring the interval between exposure and detection, using systematic colonization screening to uncover cryptic links and enable earlier recognition of transmission events.

Applying this approach, we identified 141 total transmission pairs with spatiotemporal links, representing the subset of the 361 genomically inferred transmission events with epidemiologic support for which an earliest detectable isolate pair (i.e., the first pair that could have revealed transmission) could be identified. Only 36 pairs (6%) were detected in real time. For each transmission, the first detectable isolate pair was used as the unit of analysis, representing the first opportunity to intervene in a transmission and avoid double-counting within transmission chains. Detection correlated strongly with sampling depth: none were identified from blood cultures alone; blood plus other clinical cultures yielded few (4/146, 3%); and nearly all required colonization screening, with colonization plus clinical isolates accounting for nearly all (142/146, 97%). All real-time detections involved colonization cultures. Thus, clinical isolates alone fail to capture transmission, which is reliably detected only through colonization sampling.

### Characteristics of Patients Involved in Transmission Events

A minority of patients with *S. aureus* detected in screening or clinical cultures were involved in transmissions: 46 (1.3%) in real-time epidemiologically linked events, 185 (5.2%) in delayed events, and 234 (6.6%) in genetic-only events without identifiable epidemiological links. The majority of *S. aureus* isolates (3,085/3,550; 87%) were unlinked (Table 1). Because patients could contribute to multiple events, each patient was counted once, prioritizing real-time involvement.

To determine the clinical profile of patients involved in transmission, we compared patients in epidemiologically linked events with those whose isolates were not linked to others (Table 2). Patients involved in transmission were often male (62% vs 59%), older (mean 71 vs 65 years), and more likely to have renal disease (17% vs 11.5%). They had more hospital days (34.8 vs 16), more days of antibiotic therapy (12.2 vs 3), and a higher burden of comorbidities (≥3: 22% vs 14%). Patterns were consistent across transmission groups, with transmission events identified after a delay showing greater overall healthcare exposure than those detected in real time, consistent with repeated healthcare contact contributing to delayed detection, as well as differences in sampling and time at risk. Overall, patients involved in transmission were older, sicker, and more heavily exposed to healthcare than those whose isolates were unlinked or only genetically linked.

**Table 2.**
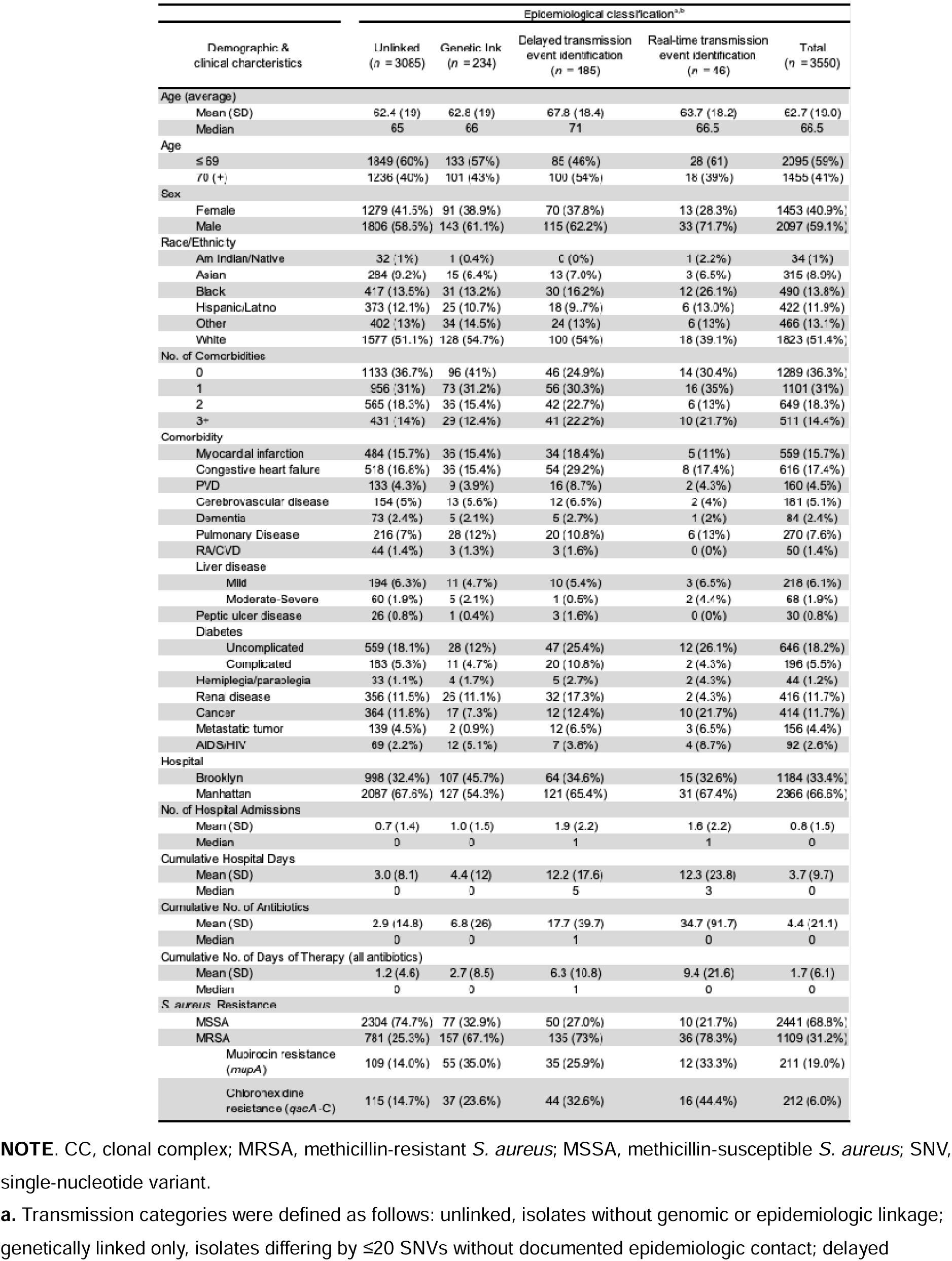
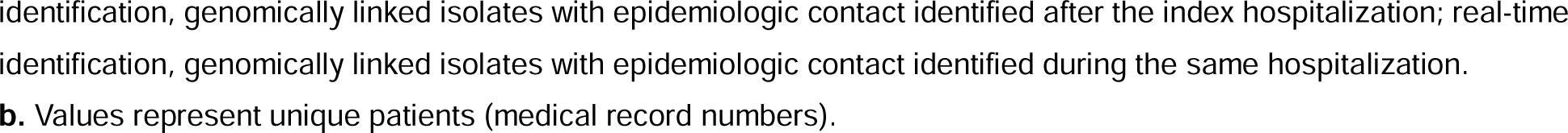
Demographic and clinical characteristics of patients involved in transmission events detected in real time. **NOTE**. CC, clonal complex; MRSA, methicillin-resistant *S. aureus*; MSSA, methicillin-susceptible *S. aureus*; SNV, single-nucleotide variant. **a.** Transmission categories were defined as follows: unlinked, isolates without genomic or epidemiologic linkage; genetically linked only, isolates differing by ≤20 SNVs without documented epidemiologic contact; delayed identification, genomically linked isolates with epidemiologic contact identified after the index hospitalization; real-time identification, genomically linked isolates with epidemiologic contact identified during the same hospitalization. **b.** Values represent unique patients (medical record numbers).

### Host–Strain Alignment and Transmission

Although MRSA transmission was dominated numerically by the CC8 lineage, transmission occurred in patients with greater healthcare exposure and comorbidity. We therefore asked whether transmission reflects alignment between host phenotype and strain lineage, specifically whether hospital-associated CC5 strains show a stronger dependence on healthcare exposure than other lineages. To test this, we modeled transmission as a function of a composite healthcare dependence (HD) score (Methods).

Figure 2 shows estimated transmission probabilities according to HD score percentiles, stratified by strain lineage. Two first-order patterns were evident: transmission probability increased monotonically with HD for all strains, and MRSA strains were more likely to transmit than MSSA across the full HD spectrum; MRSA in the lowest HD strata transmits more than MSSA in the highest HD strata. Consistent with a hospital-adapted niche, the increase in transmission with HD was most pronounced for CC5. Score tests supported a steeper HD–transmission relationship for CC5 compared with MSSA (*p* = 0.007), but not compared with CC8 (*p* = 0.40). This synergy is not apparent in aggregate transmission counts, where CC8 predominates, but emerges when transmission is examined across gradients of healthcare exposure: transmission increases with HD for both MRSA lineages, with a steeper rise for CC5 at higher percentiles, consistent with its alignment with highly healthcare-dependent hosts.

**Figure 2.**
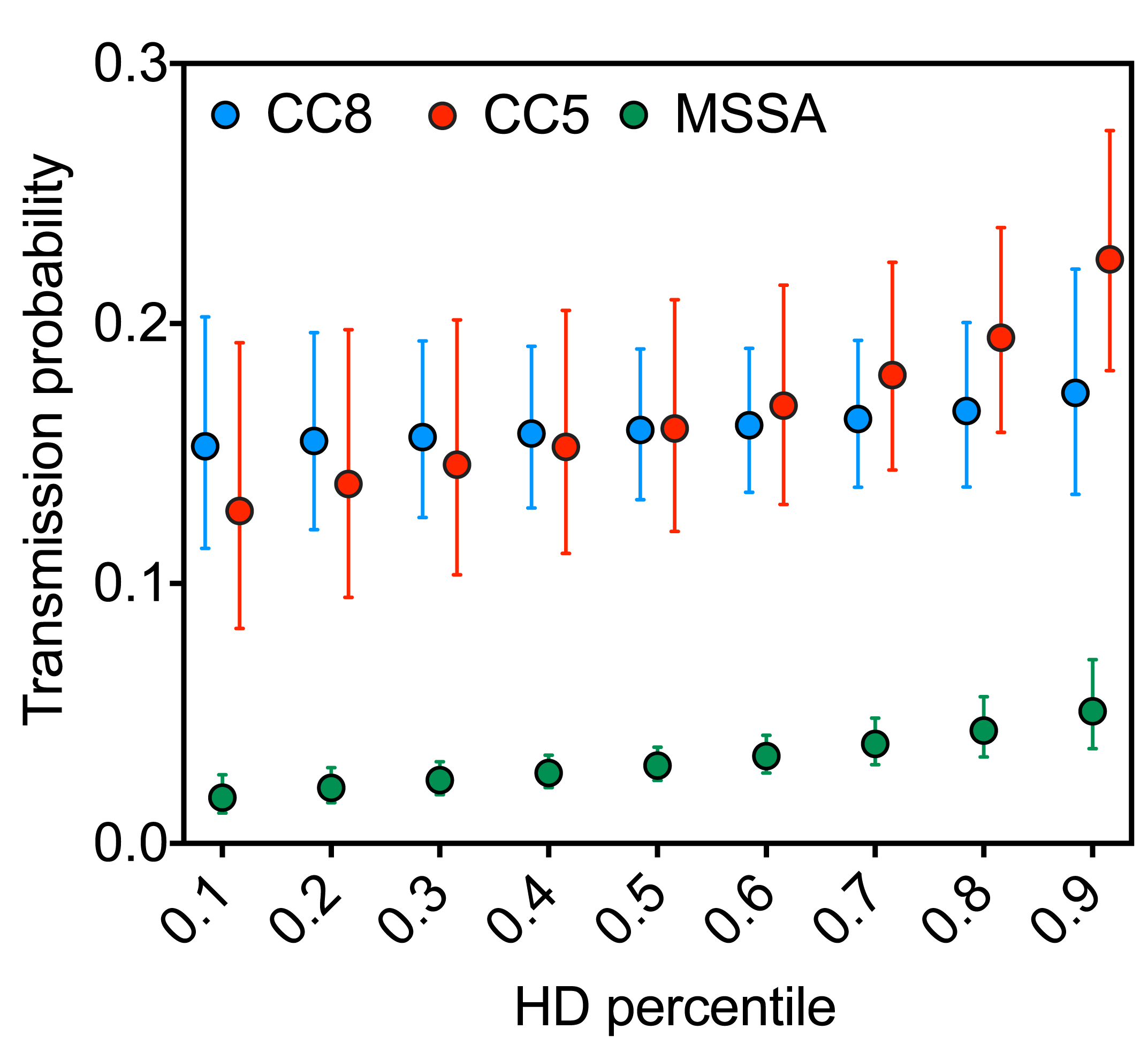
Transmission probability by strain lineage and healthcare dependence. Points show predicted transmission probabilities with 95% confidence intervals across HD percentiles for MRSA CC5, MRSA CC8, and MSSA. HD was derived from a logistic model of CC5 vs MSSA using demographic and clinical variables (Methods). Transmission was defined by genomic relatedness (≤20 SNVs) and epidemiologic linkage (direct or indirect ward contact). Host–pathogen alignment was defined as concordance between host phenotype (healthcare exposure and comorbidity, captured by the HD score; see Methods) and strain niche (hospital-associated CC5 [USA100/USA800]; community-associated CC8 [USA300]). Models were adjusted for overlap days, ward prevalence, sampling intensity, antibiotic/decolonization exposure, hospital site, ward, and calendar week (mixed-effects logistic regression). CC, clonal complex; MRSA, methicillin-resistant *S. aureus*; MSSA, methicillin-susceptible *S. aureus*.

## DISCUSSION

The present work shows that clinical isolates alone capture few transmission events. Incorporating colonization cultures increased transmission detection from 3% (2/141) to 97% (139/141), indicating that nearly all events required colonization screening for detection. Repeated sampling was also essential, since patients often carried multiple or changing strains such that a single isolate failed to capture the transmitted strain. These findings help explain why studies limited to clinical isolates often report little or no intrahospital transmission (e.g., [27]). Across all admissions, despite active screening, 72% of transmissions were detected only at readmission, indicating that spread is frequently recognized too late for intervention. Improving real-time detection will likely require sampling before discharge and, potentially, at serial time points.

Universal weekly or discharge screening for all patients would maximize detection but is unlikely to be practical or cost-effective. A more feasible strategy is targeted surveillance guided by patient- and strain-specific risk factors. Patients linked to transmissions were older, more often male, with higher comorbidity, longer cumulative stays, and increased antibiotic exposure, features overlapping with known MRSA infection risk factors. Additionally, transmission risk was amplified when host phenotype aligned with strain niche: MRSA lineages showed increasing transmission with healthcare exposure, with a steeper rise for hospital-associated CC5 (vs MSSA), a pattern not apparent in aggregate transmission counts. Together, these findings suggest that prevention strategies should target patients and strains jointly, prioritizing high-risk hosts carrying high-transmission lineages. Such features can be used to prioritize sampling and sequencing, improving efficiency while lowering cost. Whether attributes of asymptomatic transmitters also define infectious individuals remains unknown but is critical for targeting interventions.

Routine decolonization with chlorhexidine and mupirocin, widely used in our hospital, may reduce infection risk; however, our findings demonstrate ongoing transmission despite widespread use. Moreover, broad application of topical antimicrobials risks selecting for resistance. Prior work supports the idea that high-risk host populations accelerate resistance evolution [28, 29], often detected only after widespread dissemination. In contrast, real-time genomic cluster detection could direct targeted interventions, limit resistance, and concentrate resources where they are most effective.

Our study has several limitations. First, we sequenced all clinical MRSA and screening isolates but not all clinical MSSA isolates due to high volume, potentially underestimating MSSA transmission. Second, many genetically related isolates lacked clear spatiotemporal links, likely reflecting missed transmissions due to unsampled sites, healthcare workers, undetected colonization, community-based spread, or false-negative cultures. Conversely, some links may be false positives due to multiple potential contact chains or unknown sources, potentially leading to overestimation. Third, the study was descriptive: associations between comorbidities and transmission may reflect longer hospital stays, greater healthcare exposure, and increased culture frequency among sicker patients, creating more opportunities for both transmission and detection.

In sum, several implications for surveillance and intervention emerge. Because universal intensive sampling is likely infeasible, identifying patient- and strain-specific risk factors will be critical for targeted intervention. Our analyses suggest that host-strain interaction represents a complementary axis of risk stratification, providing information beyond marginal patient or strain risk alone and potentially informing efforts to distinguish transmission events more likely to result in infection. Our results provide a framework for precision genomic surveillance and lay the groundwork for trials testing whether targeted interventions can cost-effectively reduce MRSA transmission and its clinical consequences. Future work should address undetected colonization in genetically linked isolates without epidemiologic connections, for example using modeling frameworks [30, 31], and it should incorporate data on *S. aureus* introduction from the community.

## Notes

### Author contributions

K.R., Conceptualization, Data curation, Formal analysis, Investigation, Visualization, Methodology, Writing - original draft and review and editing

A.T., Investigation, Writing - review and editing

C.T., Data curation, Formal analysis, Visualization, Methodology, Writing - review and editing

M.P., Investigation, Writing - review and editing

N.S., Investigation, Writing - review and editing

J.S., Investigation, Data curation

N.A., Investigation, Data curation

A. S., Investigation, Data curation

V. T., Funding acquisition

G.P., Methodology, Data curation, Writing - review and editing

A.P., Supervision, Visualization, Writing -review and editing

S.H., Supervision, Funding acquisition, Writing – review and editing

A. R, Supervision, Funding acquisition, Project Administration, Writing –review and editing

B.S., Conceptualization, Resources, Supervision, Funding acquisition, Project Administration, Writing – original draft and review and editing

## Disclaimer

The funders had no role in the study design, data collection, data analysis, data interpretation, or writing of this report. The opinions, findings and conclusions expressed in this manuscript reflect those of the authors alone.

## Data Availability

Sequencing data are available through the NCBI repository using the accession number PRJNA1442056.

## Acknowledgments

We thank Marc Lipsitch for critical comments on the manuscript.

## Financial support

This work was supported in part by National Institutes of Health grants AI137336 and AI140754 (B.S. and V.J.T.); AI193629 (B.S. and A.R); Centers for Disease Control and Prevention grant U01CK000590 (B.S.); AI149350 (V.J.T.); and funds from the NYU Langone Health Antimicrobial-Resistant Pathogens Program (B.S., S.H., V.J.T.).

## Potential conflicts of interest

B. S. has served on a scientific advisory board for Innoviva Specialty Therapeutics and has received research funding from Analog Devices Inc. All other authors declare no competing interests. All authors have submitted the ICMJE Form for Disclosure of Potential Conflicts of Interest. Conflicts that the editors consider relevant to the content of the manuscript have been disclosed.

## Mention of any meeting(s) where the information has previously been presented

This work was presented in part at the Society for Healthcare Epidemiology of America (SHEA) Spring Conference, 2025.

## Notes

### Competing Interest Statement

The authors have declared no competing interest.

### Author Declarations

The NYU Grossman School of Medicine IRB reviewed and approved a request to waive the individual authorization requirements under HIPAA for the use and disclosure of PHI for Research under 45 CFR 164.512(i)(2)(ii)(A-C).

