## Supplementary information text (Methods), and Figures (S1 to S3) for "Sampling and host-strain interactions shape detection of *Staphylococcus aureus* transmission in hospitals"

**This PDF file includes:**

**Supplementary Information text (Methods)**

**Figures (S1 to S3)**

Fig. S1. Representative patterns of real-time and delayed detection of *S. aureus* transmission events
Fig. S2. Distribution of SNV distances among epidemiologically linked and unlinked individuals
Fig. S3. Diversity between isolates from the same individual collected on different days

**References for SI citations**

### Supplementary Information Text (Methods)

#### Decolonization Protocol and Compliance

Patients colonized with MSSA or MRSA identified from surveillance cultures were prescribed a five-day decolonization regimen consisting of daily 2% chlorohexidine gluconate (CHG) wipes applied to intact skin below the chin (excluding genitals) and mupirocin ointment applied twice daily to both nares. Patients with central venous catheters received daily 2% CHG wipes for the duration of catheter placement, regardless of *S. aureus* colonization. Compliance with admission *S. aureus* screening was 77.8%. Among patients hospitalized for ≥3 days, compliance with ≥3 days of CHG and mupirocin decolonization was 34.7%; among those hospitalized for >5 days, compliance was 56%.

#### Genomic analyses

*Genotyping and feature classification*. Sequence types and clonal complexes were assigned using MLST v2.19.0 (https://github.com/tseemann/mlst). Methicillin resistance (via *mecA* detection) and and SCC*mec* type were determined using staphopia-sccmec v1.0.0 (https://github.com/staphopia/staphopia-sccmec), part of the Staphopia pipeline (1). Clinical laboratory classification used Vitek 2 (BioMérieux) for clinical isolates and CHROMagar MRSA II (BD Diagnostics) for surveillance cultures; discordant results were resolved by genomic classification. In total, 110 colonizing isolates were relabeled based on *mecA* status, which was concordant with repeat chromogenic testing. Panton–Valentine leukocidin (*lukS/F*) and ACME (*arcA*, *opp3A*) were detected using ARIBA v2.14.6 (2), using reference sequences from FPR3757, with a minimum depth of 40× required to call gene presence.

*Variant calling and phylogenetic analysis*. Within each clonal complex, Snippy v4.6.0 (https://github.com/tseemann/snippy) alignments were merged using *snippy-core* to generate multiple sequence alignments, from which single nucleotide variant (SNV) matrices were derived using snp-dists v0.8.2 (3). Four alignments were generated: CC5-specific (JH1 reference; 2,317,809 alignment positions of which 58,487 are variable), CC8-specific (FPR3757 reference; 2,340,646 alignment positions of which 54,038 are variable), and two “all-isolate” alignments to each reference strain (JH1: 1,679,661 alignment positions of which 296,592 are variable; FPR3757: 1,678,945 alignment positions of which 296,282 are variable). Pairwise SNV distances were calculated from the most appropriate alignment: CC5-specific or CC8-specific when both isolates belonged to the same clonal complex; otherwise, the all-isolate alignment yielding the larger SNV number was used. Phylogenetic trees were generated from multiple sequence alignments using IQTREE v.2.3.6 with the GTR+F+G4 model and BIONJ for initial tree generation (4).

#### Transmission Detection

*Genomic transmission thresholds*

Transmission of *S. aureus* was estimated using a threshold of 20 single-nucleotide variants (SNVs) between patients. This threshold lies within the empirically supported 15–25 SNVs range for transmission (lower for core genome, higher for whole genome)(5, 6) and is consistent with our institutional data, in which within-host diversity and epidemiologically linked transmission pairs were almost uniformly separated by ≤20 core genome SNVs (Supplementary Figures S1-S2). To improve resolution, where available, multiple isolates per patient were analyzed to capture within-host diversity, since additional variants may bridge genomic gaps that obscure transmission links.

*S. aureus cluster detection*

Genomically linked patients were grouped into genomic clusters defined as connected components of ≥2 patients linked by at least one ≤20-SNV comparison. Cluster membership did not require all pairwise isolate distances within a cluster to meet the ≤20-SNV threshold, allowing clusters to include patients connected through intermediate hosts. Transmission events were defined at the pair level as previously described (Methods), requiring both genomic relatedness (≤20 SNVs) and epidemiologic support. Transmission clusters, distinct from genomic clusters, were defined as genomic clusters involving ≥3 patients that contained at least one epidemiologically supported transmission event. Thus, transmission events represent individual epidemiologically linked pairs, whereas transmission clusters identify larger networks of genetically related patients, consistent with in-hospital spread.

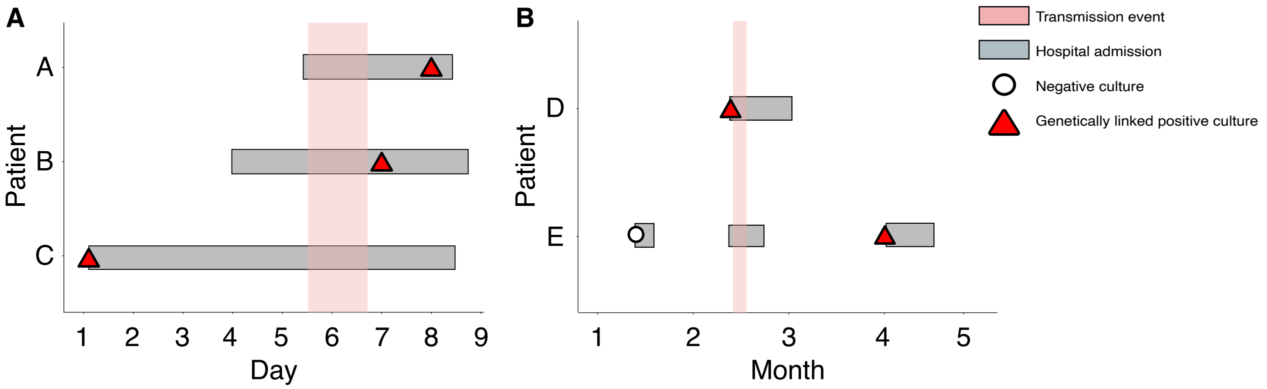


**Figure S1. Representative patterns of real-time and delayed detection of *S. aureus* transmission events.** Hospital admissions are shown as horizontal grey bars, with hatched segments indicating overlapping hospitalizations. Pink shading indicates the inferred transmission interval based on genomic linkage and overlapping admissions. Red triangles denote the first *S. aureus* isolate genetically linked (≤20 SNVs) to another isolate, and open circles indicate negative screening cultures. **(A)** Real-time detection pattern. Genetically related isolates were identified during the same hospitalization, enabling recognition of transmission during the index admission. In this example, three patients (A, B, and C) with ward contact carried genomically linked *S. aureus* isolates (≤20 SNVs), forming a transmission cluster supported by phylogenetic analysis (not shown). Patient C is the presumed source, while patients A and B are recipients. Patient B had a genetically linked clinical isolate detected while patient C was still hospitalized, whereas patient A was identified later during the same admission. The absence of negative screening cultures limits confirmation of the precise timing of acquisition. **(B)** Delayed detection pattern. Genetically related isolates were identified only after a subsequent admission. Patient E had a negative MRSA screening culture prior to ward admission which, together with phylogenetic analysis (not shown), suggests that patient D was the likely source of transmission. However, the event was not recognized during the overlapping admission (month 2) because patient E was not sampled after the period of contact. Instead, the transmission was detected later when a colonizing isolate was obtained from patient E during screening approximately two months later, during a subsequent admission (month 4).

### Figures (S1 to S3)

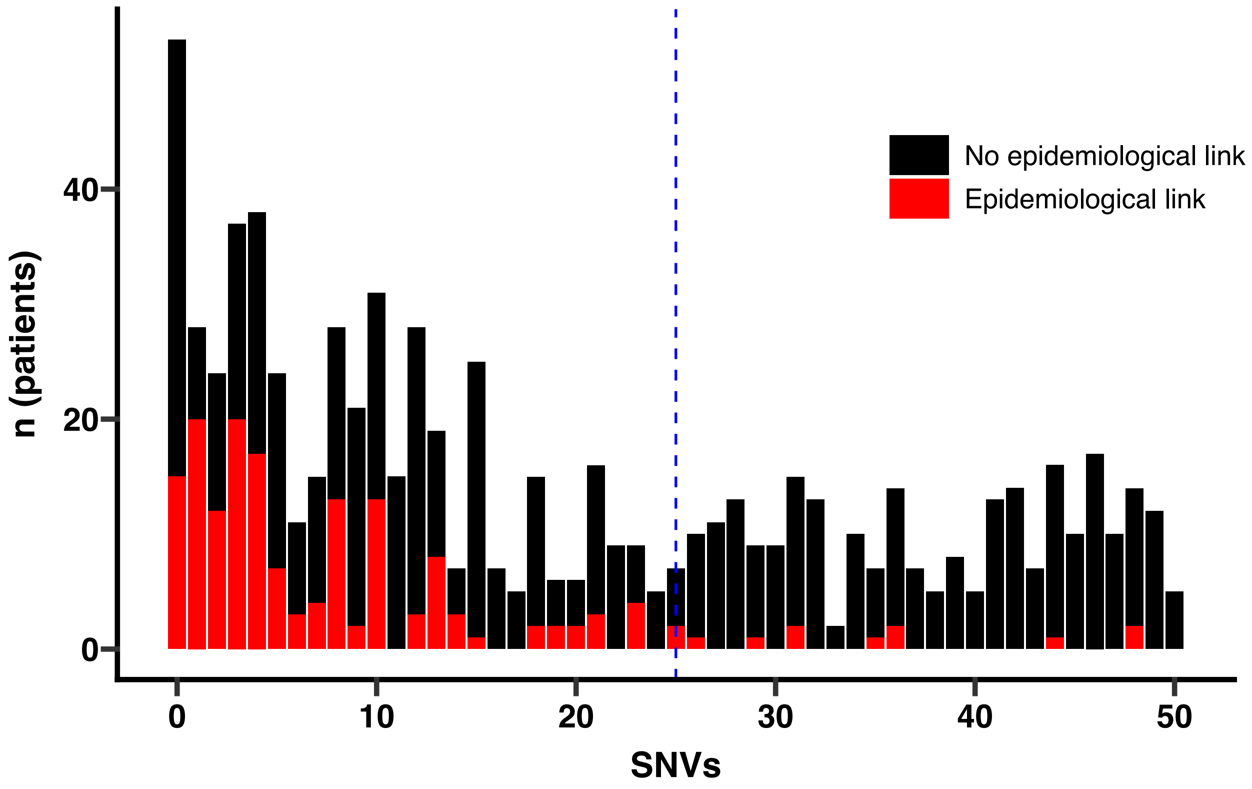


**Figure S2. Distribution of SNV distances among epidemiologically linked and unlinked individuals**. The number of patients with an *S. aureus* isolate genetically linked to that of another patient is shown as a function of the core-genome SNV (single nucleotide variant) distance. Patients are grouped by increasing pairwise SNV differences to the most similar isolate from another patient (*x*-axis), with counts shown on the *y*-axis and color-coded by epidemiologic linkage (red, linked; black, unlinked; *n* = 745). The vertical dashed blue line indicates 25 SNVs, the threshold within which 95% of epidemiologically linked isolate pairs are observed. SNV, single nucleotide variant.

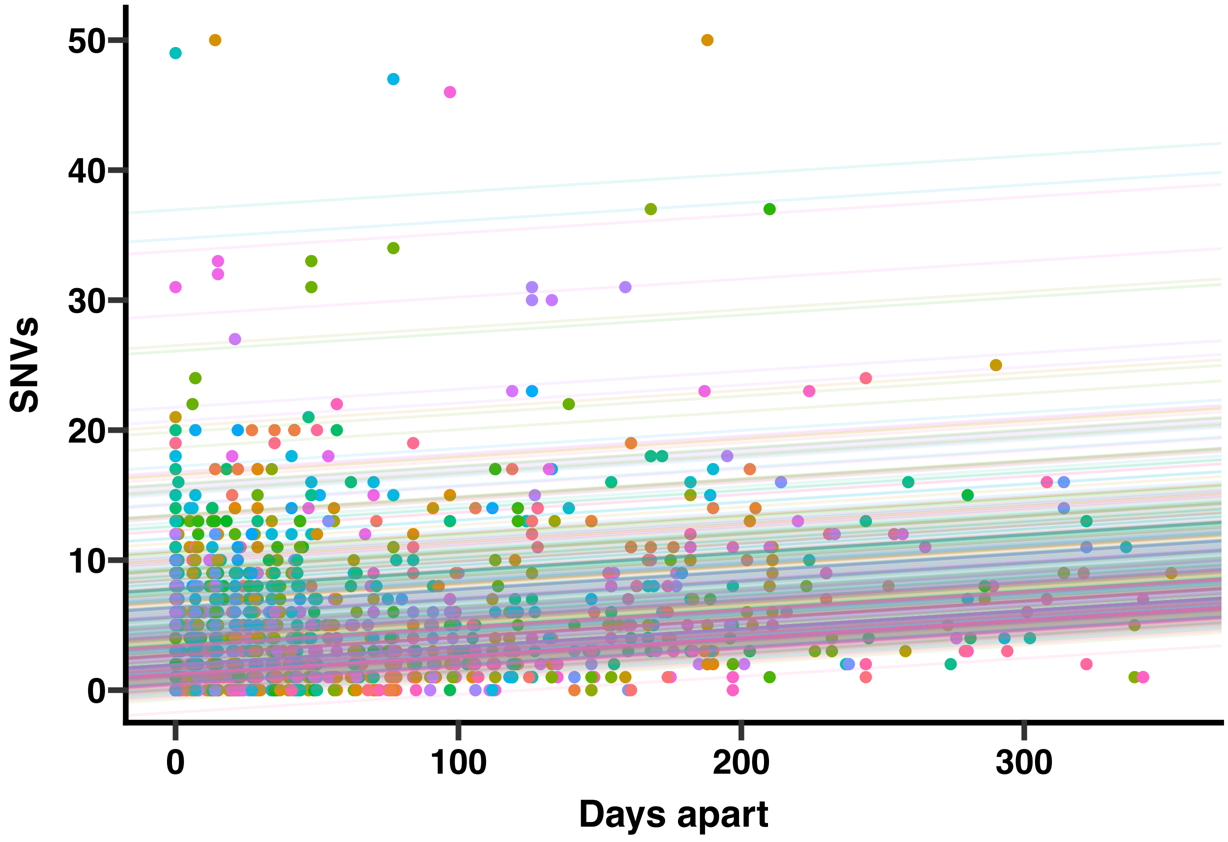


**Figure S3. Within-host genetic diversity and accumulation of SNVs over time in individual patients.** Within-host diversity of 1,713 clonally related (≤ 50 core genome SNVs) isolates from 592 patients in our collection. Each datapoint represents the genetic distance (number of core genome SNVs) between two isolates from the same individual, sampled from different sites and/or time points. Lines represent SNV accumulation over time, estimated using a linear mixed-effects regression model (R package *lme4* (7)), with SNV accumulation rate (per day) as a fixed effect and a subject-specific random intercept capturing baseline within-host diversity. SNV, single nucleotide variant. Within-host diversity, the SNV differences observed among isolates from a single patient, provides an empirical estimate of short-term evolutionary divergence during colonization or infection. This serves as an approximate upper bound on genetic distances expected among closely related isolates in a transmission chain; distances above this range are less likely to reflect recent shared ancestry (5, 6).
